# COVID-19 scenario modelling for the mitigation of capacity-dependent deaths in intensive care: computer simulation study

**DOI:** 10.1101/2020.04.02.20050898

**Authors:** Richard M Wood, Christopher J McWilliams, Matthew J Thomas, Christopher P Bourdeaux, Christos Vasilakis

## Abstract

**Background:** Managing healthcare demand and capacity is especially difficult in the context of the COVID-19 pandemic, where limited intensive care resources can be overwhelmed by a large number of cases requiring admission in a short space of time. If patients are unable to access this specialist resource, then death is a likely outcome. The aim of this study is to estimate the extent to which such capacity-dependent deaths can be mitigated through demand-side initiatives involving non-pharmaceutical interventions and supply-side measures to increase surge capacity or reduce length of stay.

**Methods:** A stochastic discrete event simulation model is developed to represent the key dynamics of the intensive care admissions process for COVID-19 patients. Model inputs are aligned to levers available to planners with key outputs including duration of time at maximum capacity (to inform workforce requirements), peak daily deaths (for mortuary planning), and total deaths (as an ultimate marker of intervention efficacy). The model - freely available - is applied to the COVID-19 response at a large hospital in England for which the effect of a number of possible interventions are simulated.

**Results:** Capacity-dependent deaths are closely associated with both the nature and effectiveness of non-pharmaceutical interventions and availability of intensive care beds. For the hospital considered, results suggest that capacity-dependent deaths can be reduced five-fold through a combination of isolation policies, a doubling of bed capacity, and 25% reduced length of stay.

**Conclusions:** Without treatment or vaccination there is little that can be done to reduce deaths occurring when patients have otherwise been treated in the most appropriate hospital setting. Healthcare planners should therefore focus on minimising the capacity-dependent deaths that are within their influence.

## 1. Introduction

Coronavirus disease 2019 (COVID-19) is a highly contagious and virulent infectious disease caused by severe acute respiratory syndrome coronavirus 2, otherwise known as SARS-CoV-2 (Anderson et al, 2020). Given the speed at which the virus can infect populations and the severity of the resulting symptoms, it represents a significant and unprecedented challenge for many healthcare services; and one with which even the most developed countries have struggled to cope (Instituto Superiore Di Sanita, 2020).

Managing a co-ordinated response to pandemics such as COVID-19 is critical. Unchecked, with a basic reproduction rate (*R0*) estimated at between 2 and 3 (Ferguson et al, 2020, Liu et al, 2020) and an estimated 4.4% of those infected requiring hospitalisation (Ferguson et al, 2020), the virus can propagate rapidly through a population (Guan et al, 2020), leading to peaks in demand for hospital care which are simply not possible to match with existing or otherwise available capacity (Ferguson et al, 2020, Instituto Superiore Di Sanita, 2020). If, at such times, patients are unable to access the bedded care required then otherwise-avoidable death is likely to result (White & Lo, 2020). The likelihood of this is particularly heightened when intensive care beds are required, since the necessary invasive ventilation and organ support cannot readily or safely be delivered in other settings (Ñamendys-Silva, 2020). Early case fatality rates from Wuhan would not be expected to appreciate these *capacity-dependent deaths* (i.e. deaths that can be attributed to a patient unable to access the care they need due to lack of available capacity), since drastic efforts were taken by authorities to avoid health services becoming overwhelmed, in enforcing restrictions on movement and rapidly upscaling capacity through the building of two new hospitals (Khan et al, 2020). Without improved treatment options, there is little that can be done to reduce COVID-19 deaths occurring when the patient has otherwise been cared for in the most appropriate hospital setting, and so planners should focus on keeping to a minimum the capacity-dependent deaths that are within their influence. That is, efforts should be made to ensure the right level of care is available to patients at the right time.

The principal levers to reduce capacity-dependent deaths relate to managing the demand for and supply of healthcare resources. On the demand side, in absence of the means to treat or prevent disease, the slowing down of cases requiring hospitalisation using measures such as school closures and social distancing can reduce peak excess demand, the so-called “flattening the curve” (Anderson et al, 2020). On the supply side, efforts to expand the capacity of medical and ventilated beds can increase the effective operating throughput, meaning more COVID-19 patients could be admitted and fewer would be rejected with either no care or care in a sub-optimal setting, resulting in increased chance of death.

The ability to use a mathematical or computer model to experiment with “what if” scenarios involving these levers is crucial to planners on the ground, in ensuring deaths over the course of the pandemic can be kept at a minimum. Public health authorities need to know what effect their policies on social distancing, home isolation and school closures (i.e. policies to reduce the effective *R0*) can have on decreasing or changing the shape over time of demand and, in turn, capacity-dependent deaths. Healthcare service planners and managers need to be cognisant of the likely benefits of their options around the flexing of bedded capacity, especially regarding the allocation between acute and intensive care beds (where the substantial efforts involved in increasing the latter must be well justified). With an appropriate model and accompanying software tool, the effect of these scenarios can be projected and used to make better informed strategic decisions when planning the response to a COVID-19 pandemic.

There has been much interest in the quantitative and mathematical modelling of COVID-19 for purposes of epidemiological forecasting (Ferguson et al, 2020, Kucharski, 2020, Roosa et al, 2020), risk prediction (Vihinen, 2020), and health system vulnerability (Gilbert et al, 2020). However, to the best of the authors’ knowledge there has been no explicit modelling of capacity-dependent deaths based on predicted demand. While Ferguson et al (2020) provide a detailed model of demand and the resulting deaths under various mitigation strategies, their work assumes a fixed mortality rate that is not dependent on the available capacity of the healthcare system. The modelling in this paper addresses this limitation by predicting the excess mortality resulting from demand exceeding intensive care capacity under several mitigation scenarios.

Computer simulations of patient flow, demand and capacity have been used extensively to inform decision-making in healthcare (Fone et al, 2003, Griffiths et al, 2013, Mohiuddin et al, 2017, Wood & Murch, 2019). This is especially true for the stochastic, discrete-event approach to simulation, as it is particularly suited to situations where entities (e.g. patients) “compete” for limited resources such as hospital beds and operating room time (Pitt et al, 2016). Many simulation studies that have tackled questions around demand and capacity in healthcare, both under typical health system conditions (for example Bagust et al, 1999, Demir et al, 2015) and in periods of increased pressure such as mass casualty events (Glasgow et al, 2018) and winter bed crises (Vasilakis & El-Darzi, 2001, Wood, 2019). Specifically in the context of intensive care, simulation studies have addressed bed requirements by using the system dynamics simulation approach to evaluate different management policies (Mahmoudian-Dehkrodi & Sadat, 2016), and applying analytical queuing models and simulations to the management of patient flow (Kim et al, 1999, Griffiths et al, 2010).

This paper reports on the development and application of a purpose-built computer simulation model and accompanying easy-to-use open source software, designed for evaluating scenarios to mitigate capacity-dependent deaths from a COVID-19 (or other) pandemic. It should be noted that capacity-independent deaths, occurring when patients are cared for in the most appropriate setting, are out of scope of this study. The remainder of this paper is structured as follows. Development of the model and implementation is covered in Section 2 alongside data requirements for model parameterisation and the scenarios considered for experimenting with the model. Illustrative results, on application to an intensive care unit in a large teaching hospital in England, are presented in Section 3. Finally, Section 4 contains a discussion on limitations, practical application, and further uses of the model and tool.

## 2. Materials and methods

### 2.1 Model and solution

The COVID-19 hospital admission process is modelled as a multi-channel queuing system operating with loss. That is, patients requiring hospitalisation are rejected if there is no available service channel (bed). In Kendall’s notation (Kendall, 1953) this is an *M*(*t*) |*G*|*C*|*C*queuing system: that is, in turn, a time-inhomogeneous Poisson arrivals process representing the *epidemic curve* for cases requiring hospitalisation; a general service distribution approximating patient length of stay in hospital; *C* service channels; and a total system capacity of *C* patients, i.e. no space for waiting. For rejected admissions (lost arrivals), death occurs with probability *P*_*d*_ and survival with probability 1−*P*_*d*_. The model can be applied in the context of general acute beds or intensive care beds, assuming the parameters are calibrated accordingly.

Implementation of this model is through the iterative three-phased method of discrete event simulation (Pidd, 1998). In the case of this study, the types of simulation event consist of:

a. Arrival of patient requiring hospital admission (unconditional event)
b. Patient admitted (conditional event)
c. Patient discharged (unconditional event)
d. Patient admission rejected and patient died (conditional event)
e. Patient admission rejected and patient survived (conditional event)

It should be noted that event type c includes the quite substantial potential (e.g. 61.5% of patients admitted to ICU had died within 28 days, Yang et al, 2020) for patients to die within hospital. This should be considered a *capacity-independent death*, and thus it is not within scope of this paper.

The basis of the three-phased approach is in maintaining a calendar of unconditional events. The first phase is to step to the next chronological event in the calendar. This could be arrival or discharge (i.e. event type a or c as above). In the second phase this event is executed. The third phase sees any associated conditional event also executed. So, for example, if a patient arrives (event type a) and there is an available service channel (e.g. a free bed) then the conditional event is that the patient is admitted (event type b) and the bed is flagged as unavailable. If, instead, there is no available service channel then their admission is rejected and he or she either dies (event type d) or ultimately survives (event type e).

As the simulated events progress with each iteration, it is necessary to capture the state of the system over time. This keeps the event calendar up-to-date. For instance, if one of the events within an iteration involves a patient entering service (event type b) then the time at which they are discharged (sampled from the given length of stay distribution) is recorded in the calendar, as a future unconditional event (of type c). Capturing the state of the system is also necessary in the generation of performance measures of interest, such as occupancy levels and patient outcomes.

During the simulation, events are iterated in line with the three-phased method until some terminating criterion is met. Here, this is given by the time at which some outcome has been reached for all simulated admissions for the given epidemic curve (for cases requiring hospitalisation), i.e. each sought admission has been either rejected or admitted and discharged (event types c-e). In other words, and given the time-inhomogeneous nature of the epidemic curve, this is a transient simulation model. As such, and in contrast to simulation models exploring steady-state behaviour, an otherwise necessary warm-up period is not required (Law & Kelton, 2000).

Running this simulation from start to finish offers just one possible explanation of how the pathway dynamics can play out and so, in order to capture the inherent stochasticity, it is necessary to perform an ensemble of replications. Each replication repeats the simulation with a different stream of random numbers from which the simulated arrivals, lengths of stay, and rejection probabilities of death and survival are generated. Outputs are then aggregated across these replications, with central estimates (based on the mean), inter-quartile range (IQR), and confidence bands (at the 95% level) calculated for the performance measures considered.

### 2.2 Application, data, and calibration

The model is applied to intensive care services at a major public hospital in England. Following a similar approach to Deasy et al (2020), demand for intensive care admission is estimated through local interpretation of nationwide projections contained in Ferguson et al, 2020 (controlling for local population size, demographics and hospital catchment area – see Table 1). This is according to two scenarios, as presented in Ferguson et al (2020). The first is effectively a “*do nothing*” involving no restrictions on movement, while the second involves “*case isolation, home quarantine, and social distancing of those over 70*” (Figure 1). Informed by publication of this report on 16^th^ March 2020, the UK Government implemented restrictions on movement equivalent to this latter strategy, effective as of 20^th^ March 2020.

**Table 1.** Distribution of ages within estimated hospital catchment area.

| Age bands | Proportion of hospital catchment |
| --- | --- |
| 0-9 | 11% |
| 10-19 | 10% |
| 20-29 | 21% |
| 30-39 | 15% |
| 40-49 | 10% |
| 50-59 | 10% |
| 60-69 | 7% |
| 70-79 | 6% |
| 80+ | 11% |

**Figure 1.**
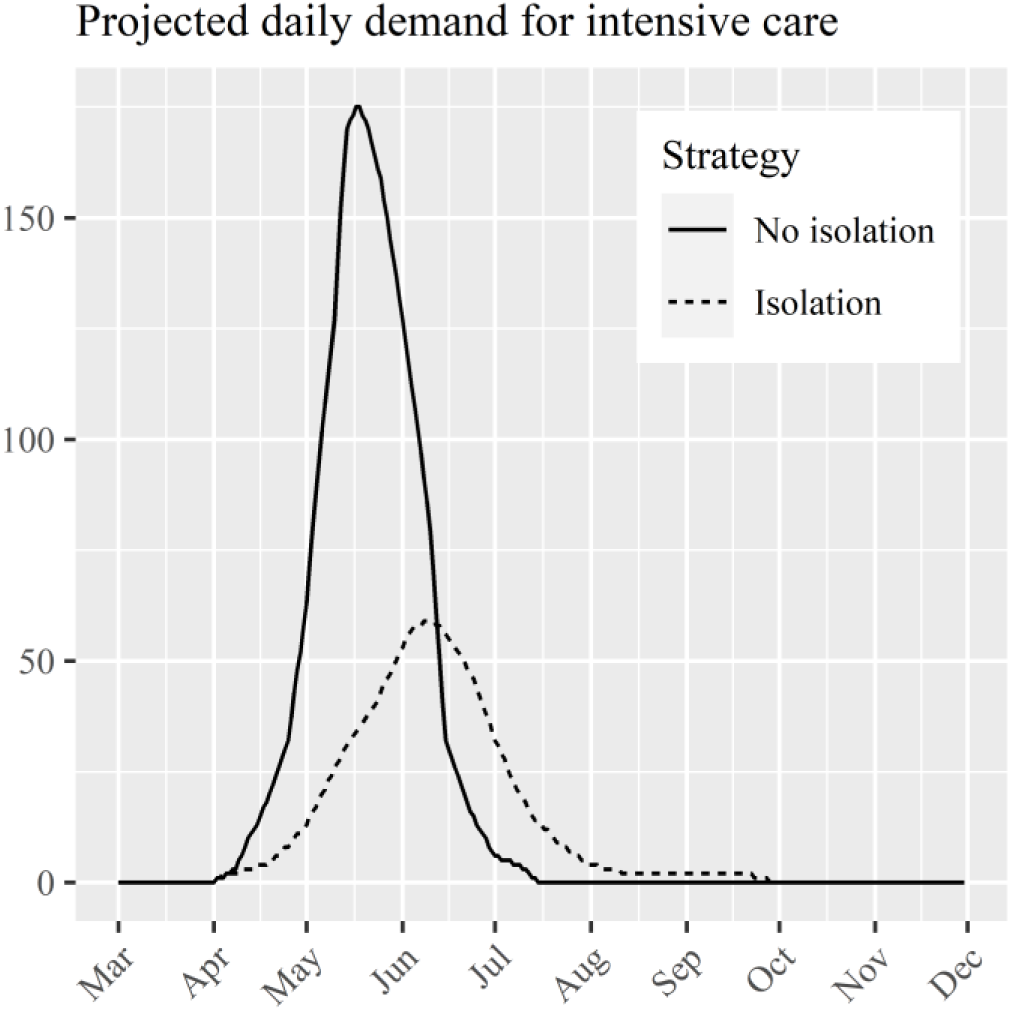
Epidemic curve for cases requiring intensive care, derived from modelling results in Ferguson et al (2020). The *No isolation* scenario assumes no non-pharmaceutical intervention; *Isolation* assumes case isolation, home quarantine, and social distancing of those over 70.

At the hospital there are typically 45 beds available for patients requiring intensive care (21 general and 24 cardiac). In the first instance, plans are in place for capacity to be increased to a maximum of 76 beds, through making use of operating theatres and other specialist bays (which have become available due to the cancellation of routine surgery). There remains some potential to increase this number further, should additional surge capacity be required (this is considered within the scenario analysis of Section 3).

Given that only a handful of COVID-19 patients are, at the time of the study, admitted to an intensive care bed at the hospital (and none have either died or been discharged, i.e. observations are right-censored), information regarding intensive care length of stay is taken from the literature. Deasy et al, 2020 estimate this to be gamma-distributed with *α* =8and *β*=1(see Figure 2), based on an analysis of experience from Wuhan (Zhou et al, 2020, Yang et al, 2020). Early observations of COVID-19 intensive care lengths of stay in the UK as of 27^th^ March 2020 (ICNARC, 2020) appear to be consistent with this distribution. The probability of death resulting from rejected admission to intensive care (*P*_*d*_) was also informed by the literature. Given the pivotal dependence of survival on mechanical ventilation (White & Lo, 2020) and already very high mortality rates (81-97%) for cases actually receiving such intervention (Weiss & Murdoch, 2020), it is assumed that all but a very small minority of rejected admissions would result in death. For the simulation study conducted here, a figure of *P*_*d*_=0.99is used.

**Figure 2.**
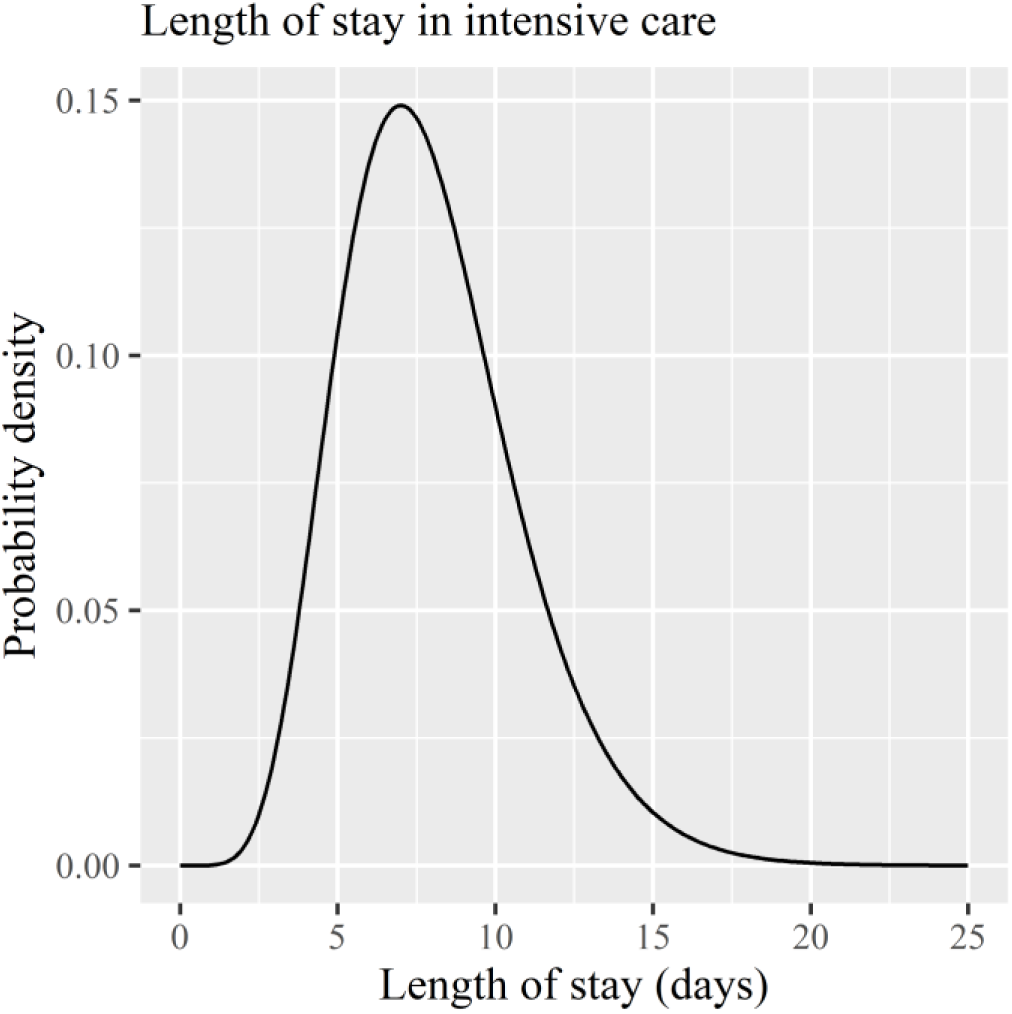
Distribution of length of stay for COVID-19 patients in intensive care, as suggested within Deasy et al, 2020.

### 2.3 Simulation scenarios

A number of scenarios relating to possible COVID-19 mitigations are modelled in order to inform planning of intensive care services at the hospital (Table 2). These relate to changes in the epidemic curve for cases requiring intensive care (informed by government-led strategy regarding isolation, quarantine and social distancing), capacity at the hospital in terms of number of intensive care beds, and the patient length of stay in intensive care. The *No isolation* strategy involving no government-led effort with respect to isolation, quarantine and social distancing is considered within Scenario 1, alongside the current available capacity of 45 beds and the literature-informed gamma-distributed length of stay with mean 8 days. Given the UK Government’s decision on 20^th^ March 2020 to implement isolative measures, the remainder of scenarios (2 through 7) are configured on the basis of this afore-mentioned *Isolation* strategy (Section 2.2).

**Table 2.** Simulation scenarios considered in this study. Note that strategies under Scenarios 1-5 relate to the epidemic curves for cases requiring intensive care equivalent to those contained in Figure 1, with Scenarios 6 and 7 based upon a 50% flattened version of the *Isolation* curve containing the same total demand but spread over a 1.5-fold longer period of time.

| Scenario | Strategy | Capacity (intensive care beds) | Mean length of stay (days) |
| --- | --- | --- | --- |
| 1 | No isolation | 45 | 8 |
| 2 | Isolation | 45 | 8 |
| 3 | Isolation | 76 | 8 |
| 4 | Isolation | 100 | 8 |
| 5 | Isolation | 45 | 6 |
| 6 | Isolation (flattened) | 45 | 8 |
| 7 | Isolation (flattened) | 100 | 6 |

Scenarios 3 and 4 model the hospital’s actual planned increases in intensive care bed numbers to surge capacities of 76 and 100 respectively. Scenario 5 models the potential benefits of reducing COVID-19 length of stay (for example by using digital interventions to optimise clinical practice and workflows e.g. Bourdeaux et al 2016, McWilliams et al 2019) by considering a one-quarter reduction in mean length of stay. There may be scope for government-led strategies to further restrict movement and thus the rate at which infections are acquired in the population. Scenarios 6 and 7 model such an eventuality by stretching the epidemic curve for cases requiring intensive care (under the *Isolation* strategy) by 50% (i.e. the same demand and shape, just spread over a 50% longer time period). The final scenario involves a “best case” option in bringing together flattened demand, increased capacity and reduced length of stay.

Key simulation output measures of interest consist of the duration of time at maximum capacity (to inform workforce requirements), peak capacity-dependent deaths per day (for mortuary planning), and total capacity-dependent deaths over the course of the pandemic (as an ultimate marker of intervention efficacy, in balancing demand and capacity). Quantiles, including inter-quartile range (IQR) and 95% confidence intervals, are calculated based on the variation in output measure observed across the 1000 replications performed for each scenario.

## 3. Results

Estimates for the key output measures of interest are presented alongside each of the considered scenarios in Table 3. Transient outputs corresponding to each of these key areas of interest are presented in Figure 3 across all scenarios. This highlights the key dynamical relationships between these variables. For instance, when full capacity is reached (left plots) then capacity-dependent deaths start to occur (middle plots) based on the extent to which demand continues to exceed supply; with the magnitude of this determining the rate at which deaths accumulate (right plots).

**Table 3.** Simulation key output measures of interest obtained over 1,000 simulation replications. Note that strategies under Scenarios 1-5 relate to the epidemic curves for cases requiring intensive care equivalent to those contained in Figure 1, with Scenarios 6 and 7 based upon a 50% flattened version of the *Isolation* curve containing the same total demand but spread over a 1.5-fold longer period of time. Note * denotes capacity-dependent deaths following rejected intensive care admission.

| Scenario | Continual days at maximum capacity; mean (IQR) | Peak daily deaths*; mean (IQR) | Total deaths* over pandemic; mean (IQR) |
| --- | --- | --- | --- |
| 1 | 67 (61-79) | 167 (154-181) | 5887 (5511-6250) |
| 2 | 68 (61-93) | 53 (47-59) | 2521 (2342-2696) |
| 3 | 62 (39-77) | 49 (43-56) | 2172 (2000-2345) |
| 4 | 57 (57-74) | 46 (39-53) | 1931 (1762-2096) |
| 5 | 59 (51-85) | 51 (45-58) | 2350 (2173-2524) |
| 6 | 82 (67-125) | 33 (28-38) | 2296 (2117-2478) |
| 7 | 31 (34-79) | 22 (17-27) | 1152 (1002-1300) |

**Figure 3.**
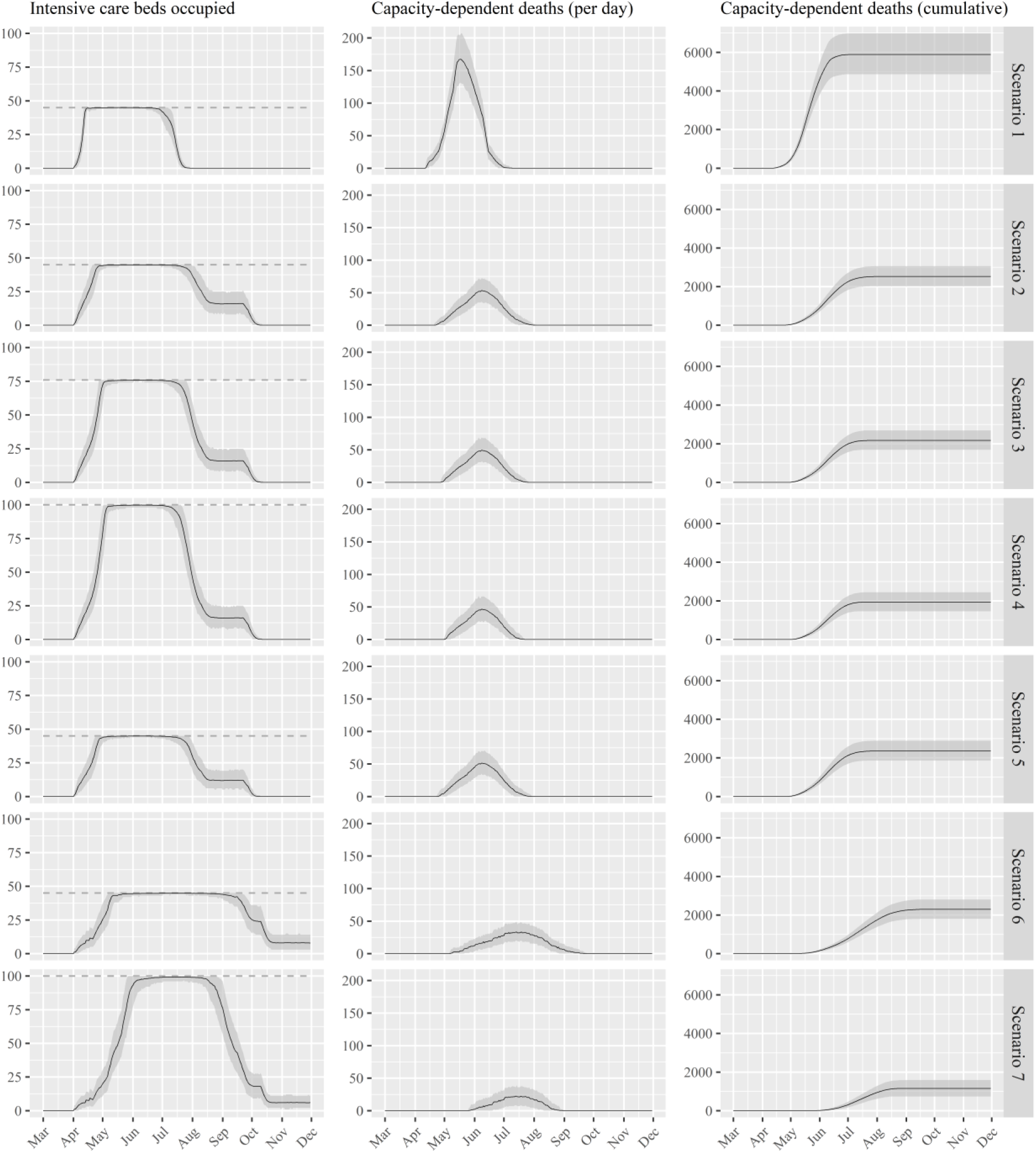
Simulation output results for the seven scenarios (see Table 2). Black solid lines represent the mean with grey bands containing the 95% confidence intervals. Dashed lines represent capacity associated with respective scenarios (provided as an input to the simulation).

In the absence of any intervention to reduce the basic reproduction rate (*R0*) through case isolation, home quarantine and social distancing (i.e. the *No isolation* strategy of Scenario 1), the estimated death toll is significantly higher than otherwise. Employing these measures reduces total deaths by an estimated 57% and cuts the peak daily deaths by more than two-thirds *ceteris paribus* (Scenario 2). Increasing capacity from 45 to 76 intensive care beds (Scenario 3) further reduces total deaths by 14%, with an effect starting to show on the number of subsequent days at maximum capacity (reducing from 68 to 62). This is brought down further (to 57 days) should capacity increases to 100 beds be possible (Scenario 4), which also brings down total deaths to under 2000. Reducing mean length of stay by one-quarter appears to have a relatively small improvement to the total number of deaths (Scenario 5 *c*.*f*. Scenario 2), which is in part due to the right-skewed nature of the length of stay distribution (i.e. the number of longer-staying patients in the tail is unchanged since the shape of the distribution is presumed unaltered).

Should any additional government-led isolation strategies be effective in further flattening the epidemic curve for cases requiring intensive care, then a substantial reduction in peak deaths from 53 to 33 would be expected (i.e. Scenario 6 *c*.*f*. Scenario 2). However, without increases to capacity this simply spreads the deaths over a longer period of time, rather than reducing the total by a significant amount (2296 *c*.*f*. 2521). To achieve a significant reduction in total deaths then any “flattening” of demand must be accompanied by increases in capacity. If this can be accomplished, alongside the afore-mentioned reductions in length of stay, then such synergy becomes evident, cutting total capacity-dependent deaths to just over 1000, reducing the peak per day to 22, and shortening the continuous time operating at full capacity.

In order to estimate the number of intensive care beds required to satisfy all demand, the model can be run without a constraint on capacity. Results, under the *Isolation* strategy with 8 days mean length of stay, are provided in Figure 4, showing a peak requirement of 466 beds (383 – 554, 95% CI).

**Figure 4.**
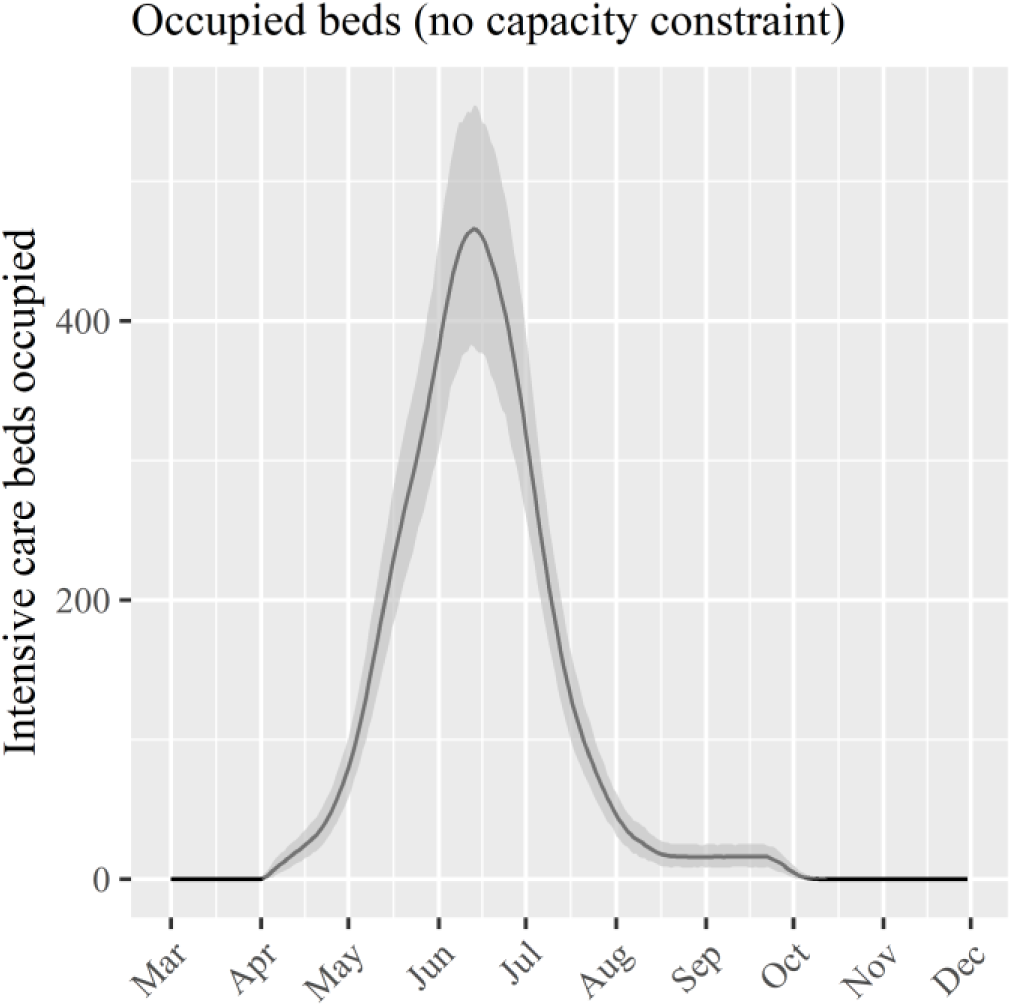
Simulation output results for no constraint to bed number availability. This shows the number of intensive care beds that would be required to satisfy all demand.

## 4. Discussion

Government and healthcare planners should focus on keeping to a minimum deaths that are within their influence, in taking measures to maximise the chances of survival by ensuring the right level of care is available when needed. As has been illustrated in this paper, such *capacity-dependent deaths* are closely associated with the nature of the healthcare response. The modelling and tool presented here offers planners a readily-accessible means to understand the responsiveness of capacity-dependent COVID-19 deaths to different demand profiles as well as plausible interventions under their control.

The modelling undertaken in this study has informed on-the-ground planning of healthcare services in response to the COVID-19 pandemic, both within the healthcare system in question and more widely through public release as an *R*-based open source tool (hosted on github.com/nhs-bnssg-analytics and promoted via social media). Results have been useful in highlighting the need for additional intensive care beds, where decision-makers must now weigh-up the benefits of converting more beds for such purpose against the opportunity cost of such actions (e.g. if theatre space is used then this may limit the ability to perform emergency surgery). Simulation results have also been useful in understanding workforce implications (through measures relating to estimated continual time at maximum occupancy) and the planning of mortuary capacity (through understanding peak daily deaths). While the number of deaths reported in this modelling study may appear to be large against the estimated 353,862 catchment area of the hospital, they should be viewed in the context of other projections. For example, the age-stratified case fatality rates of Verity et al, 2020 produce an estimated 4,349 deaths when no effort is made to explicitly model capacity-dependence in the numbers of deaths.

As with any modelling study, a number of simplifying assumptions were made. There is the assumption that death occurs immediately if a bed in the required setting is not available. Realistically death will not be immediate (World Health Organization, 2020), yet at this early stage of the pandemic there exist no reliable data to capture this parameter in the model in a meaningful way. This has no effect on the ultimate number of deaths estimated, but will affect their specific timing and the thus, the peak daily number. This should therefore be considered if seeking validation against actual number deaths over time (i.e. it should be expected that there will be a lag). It should also be acknowledged that the model does not mechanistically capture delays to discharge or transfer, which are commonplace in hospital patient flow (Landeiro et al, 2019). An example for the application considered here would be the inability to discharge a patient from intensive care due to the lack of an available acute bed. While this has not been modelled (this would be possible at the cost of additional complexity, see Wood & Murch, 2019), the effects can be understood by adjusting the length of stay distribution used within the simulation according to estimated or hypothetical delay times. Finally, it is assumed in this study that all intensive care beds are available for newly-arriving COVID-19 patients. While elective procedures requiring post-operative intensive care have been cancelled, there remains other sources of non-elective non-COVID-19 intensive care demand. Estimations of this, once the effect of societal isolation becomes appreciable (e.g. any reduced road traffic accidents, alcohol-related injuries), can be incorporated within the model parameter for capacity simply by deducting the average beds occupied by such patients.

Regarding future use of the tool and as the COVID-19 pandemic evolves and more empirical data become available, it will be possible to derive more accurate assessments of hospital length of stay and projected cases requiring hospitalisation. Given efforts to promote user-friendliness of the tool (by way of execution through a simple purpose-built function in *R*), any such changes to input parameter values can be readily addressed by running a small number of additional simulation runs (thus helping to ensure results remain in line with the latest data and forecasts). For the intensive care application considered here, the tool may also be used at a later stage of the pandemic in order to understand when elective cardiac surgery may safely resume following any projected reduction in COVID-19 intensive care occupancy. Further development of the model may also unlock the ability to assess the implication of different intensive care admission criteria on aggregate measures of mortality (as suggested in Utley et al, 2011 and White & Lo, 2020). In such case the model may be used to understand the effects of rejecting intensive care admissions from patients of cohorts known to have negligible survival likelihood, in the interests of maintaining available beds for those known to have more favourable chances.

## Data Availability

Data for this study is from publicly-available sources and made available on github.

https://github.com/nhs-bnssg-analytics/covid-simr-hospital-application

## Acknowledgements

The authors are grateful to Ben Murch for reviewing the model code used within the tool presented in this study.

